# Machine Learning Analysis of Routine EEG Accurately Predicts Anti-Seizure Medication Response

**DOI:** 10.1101/2025.10.21.25338468

**Authors:** Peter D. Galer, Gregory G. Grecco, Fan Zhang, Amulya Mathur, Jonathan J. Halford, Andrew J. Cole, Amy S. Chappell, Elizabeth Garofalo, Steven M. Paul, Michael Detke, Jacqueline A. French, Ken Wang, Qiang Li

## Abstract

Despite the availability of more than 20 anti-seizure medications (ASMs), approximately half of patients with newly diagnosed epilepsy fail their first drug trial. Unfortunately, clinicians lack objective tools or consensus guidelines to match individual patients with the most effective therapy, frequently leading to years of uncontrolled seizures. Here, we developed machine learning models to utilize a single, baseline routine resting-state scalp EEG to forecast ASM efficacy. EEGs and treatment outcomes were drawn from 280 participants with new-onset focal epilepsy in the prospective, multicenter Human Epilepsy Project. Recordings were acquired within four months either before ASM initiation (unmedicated EEG) or after treatment onset (medicated EEG). For each recording we computed band-limited static and dynamic functional-connectivity and entropy-based matrices in consecutive time windows. We trained and tested classifiers in a nested fashion to predict future seizure freedom. Separate classifiers were trained to (i) predict levetiracetam response from unmedicated EEGs (22 responders, 32 non-responders) and from EEGs recorded on an ASM (53 responders, 31 non-responders); (ii) predict lamotrigine response from unmedicated EEGs (12 responders, 21 non-responders); and (iii) distinguish participants who ultimately proved refractory to all ASMs from unmedicated EEG (67 responders, 16 refractory) and EEGs recorded while on an ASM (34 responders, 80 refractory). Two model architectures were tested for each classifier. Performance, evaluated with nested leave-one-out cross-validation, was robust across at least one model architecture for each classifier: area under the ROC curve (AUC) 0.88 and balanced accuracy 0.85 for unmedicated levetiracetam, AUC 0.82 and balanced accuracy 0.77 for medicated levetiracetam, AUC 0.79 and balanced accuracy 0.80 for unmedicated lamotrigine, AUC 0.92 with balanced accuracy 0.87 for unmedicated refractory, and AUC 0.82 with balanced accuracy 0.75 for the medicated refractory model. These findings indicate that routine EEG harbors machine-learning-detectable signatures predictive of specific ASM efficacy, laying groundwork for precision-medicine tools that could shorten the costly trial-and-error period in epilepsy treatment.

## INTRODUCTION

Epilepsy affects over three million individuals in the United States [1, 2]. Individuals living with recurrent seizures experience higher morbidity and mortality rates, decreased quality of life, and increased healthcare utilization [3-9]. In the United States, epilepsy accounts for over 1 million emergency room visits, 280,000 hospital admissions, and accounting for inflation, approximately $3.4 billion annually in hospital costs alone [10]. An outsized proportion of these admissions and healthcare costs are due to individuals with uncontrolled epilepsy, having approximately 1.7 times the annual cost of those with controlled epilepsy [10]. In addition, there are substantial indirect costs to the individual due to lost earnings, unemployment, reduced productivity, and premature mortality [5].

Despite the introduction of over 20 ASMs, including widely used medications such as levetiracetam and lamotrigine, seizure freedom rates have remained relatively unchanged over the past several decades, with nearly half of newly diagnosed epilepsy patients failing their first medication trial [11]. This lack of an objective method or consensus guidelines for initial ASM selection results in prolonged ineffective treatments, unnecessary side effects, and significant patient and healthcare system costs [12, 13], highlighting an urgent need for predictive biomarkers and algorithms to personalize epilepsy management.

Electroencephalography (EEG) is a non-invasive, widely available, and cost-effective tool for capturing real-time brain activity that is already routinely integrated into the care of every patient with epilepsy in most developed nations. Leveraging these strengths, EEG-derived biomarkers and algorithms have become a promising modality for epilepsy diagnosis [14, 15]. However, very few EEG-based biomarkers and algorithms currently exist for predicting responses to specific ASMs, instead focusing on general ASM response vs treatment resistance [16-21], limiting practical clinical utility. While some recent studies employing machine learning techniques have demonstrated feasibility in predicting individual ASM responses based on EEG recordings, such as levetiracetam [22, 23], oxcarbazepine [24], and brivaracetam [25], these efforts are often limited by modest sample sizes, risk of overfitting due to model complexity, and a lack of internal validation through nested cross-validation and external validation, raising concerns over their robustness and generalizability. Despite these limitations, these preliminary findings suggest the feasibility of developing reliable, targeted EEG-based biomarkers through machine learning approaches.

The Human Epilepsy Project (HEP) provides a robust dataset to address these limitations, consisting of EEG and clinical data from a large, multicenter prospective cohort of newly diagnosed patients with focal epilepsy [26, 27]. The HEP cohort is particularly valuable, with standardized EEG data collected at or near ASM initiation, offering an ideal framework for developing and validating predictive biomarkers suitable for real-world clinical application.

In this study, we aim to develop and validate EEG-based machine learning biomarkers that accurately predict responses to levetiracetam and lamotrigine and identify refractory epilepsy status from EEGs recorded at 28 different hospitals in the United States. Leveraging our validated computational pipeline that achieved high predictive accuracy and robustness in earlier major depressive disorder studies [28], we utilize both established and novel functional connectivity and entropy-based biomarkers extracted from baseline, routine EEG recordings to model the treatment response of patients in the HEP cohort. Importantly, our primary results are from nested cross-validated models, ensuring no data leakage and significantly increasing the chance of reproducibility. This approach explicitly addresses previous methodological limitations by utilizing concise, clinically practical EEG sessions, employing rigorous nested cross-validation strategies, and carefully selecting EEG features to minimize overfitting. The results presented here demonstrate concrete foundations for the development of robust machine learning models to accurately predict specific ASM responses in individuals with epilepsy receiving a clinical EEG.

## METHODS

### Participants and Dataset

Data for this study were obtained from the Human Epilepsy Project 1 (HEP), a prospective, multicenter observational study that enrolled individuals with newly diagnosed focal epilepsy. Participants aged 12 to 60 years at the time of seizure diagnosis were enrolled within four months of initiating anti-seizure medication (ASM) treatment and were required to have an estimated IQ of at least 70. Participants were recruited from 34 epilepsy centers across North America, Europe, and Australia between June 2012 and November 2017, with follow-up continuing until February 2020. All participants provided informed consent, and institutional review board approval was obtained at all sites. Exclusion criteria included epilepsy resulting from traumatic brain injury or other central nervous system insults, progressive neurological disorders, autism spectrum disorders, significant developmental delays or cognitive impairments (IQ<70), chronic drug or alcohol abuse, and significant psychiatric disorders interfering with study participation. Seizure history was obtained from individuals’ seizure diaries or available medical records. Complete enrollment criteria and additional methodological details are described here: [26].

To obtain sufficient sample sizes to train machine learning models, we restricted our analysis to the subset of HEP participants who: (i) had routine EEGs of sufficient technical quality and length following preprocessing, (ii) initiated treatment with levetiracetam or lamotrigine (the two most commonly prescribed first-line ASMs in this cohort) or (iii) met a clearly defined standard for treatment refractoriness, (iv) had MRI annotations that, when available, did not indicate a significant morphological abnormality, and (v) had a clear and conclusive annotated response to an ASM. Limiting the dataset in this way ensured that every subject contributed both reliable EEG features and unambiguous clinical outcome labels based on expert-derived annotations. This focus reduced heterogeneity and missing-data bias, allowing the models to learn from consistent inputs and to target clinically actionable questions relevant to everyday practice.

Treatment response was defined a priori according to International League Against Epilepsy (ILAE) criteria. An ASM responder was classified as a patient that achieved ≥12 months of seizure freedom (or a seizure-free interval at least three times longer than their longest pretreatment inter-seizure interval) [29]. To train and test the levetiracetam and lamotrigine models, their cohorts were composed of individuals that were responsive and non-responsive to the respective ASM. Non-responsive individuals were made up of both individuals that went on to respond to other ASMs and individuals that responded to no ASMs trialed and went on to be designated as treatment refractory. A patient was classified as treatment refractory if they failed adequate trials of two or more appropriately dosed, tolerated ASMs and did not respond to any subsequent ASM. This latter criterion differs from current ILAE criteria. This addition was made for two reasons. First, it is clinically important to patients and their clinicians if they will go on to achieve seizure freedom, even if it is not achieved until the third or even tenth ASM trialed. Furthermore, we hypothesized that individuals that go on to obtain seizure freedom after failing their first two ASM trials have underlying electrophysiology that more closely aligns with responder populations rather than treatment-refractory populations. To test this hypothesis and minimize potential confounders from this subpopulation, we held out individuals that were refractory by ILAE criteria but achieved seizure freedom after failing their first two adequate ASM trials. This encompasses about 4-5% of individuals in the general epilepsy population [11, 30].

### EEG Acquisition and General Preprocessing

All EEG recordings utilized for this analysis were collected in routine clinical practice according to standard protocols at each participating site. Recordings used standard 10-20 EEG electrode placement and included electrodes A1 and A2 and one EMG channel. Acquisition parameters varied slightly across sites. EEG data preprocessing was standardized using a robust pipeline based on standard practices designed to minimize artifacts and ensure consistency across recordings [14, 31]. To help ensure only resting state was captured and minimize potential differences due to varying lengths of EEG, recordings were cropped to the first four hours. Initial preprocessing was on the original recorded waveform and included removal of the A1, A2, and EMG channels, applying a notch filter at 60 Hz and its harmonics to remove electrical line noise. For EEG data sampled above 200 Hz, a low-pass filter at 95 Hz was applied to prevent signal aliasing, followed by downsampling to a uniform rate of 200 Hz. Channel labels were standardized to the traditional 10-20 system. Cleaned EEG data were then filtered using a second-order bandpass filter between 0.5–70 Hz to eliminate low-frequency drift and high-frequency noise. EEG signals were segmented into consecutive one-second non-overlapping epochs, and artifact rejection was performed using automated voltage thresholds and statistical measures such as mean line-length outliers (>3 standard deviations from the mean). The remaining consecutive clean epochs were concatenated for feature extraction. Final epoch length and overlap were tailored to the specific feature type. A minimum of two minutes of clean epochs was required for each EEG.

### Feature Extraction and Selection

Feature extraction primarily emphasized static and dynamic functional connectivity and entropy-based metrics, reflecting the known network-based pathophysiology of epilepsy. Frequency-band-specific features (delta, theta, alpha, low-beta, high-beta, and gamma) were extracted, given established associations of these oscillations with neurological dysfunction. A wide range of both established and novel features were explored and developed. Among the well-established connectivity metrics, we calculated the phase-locking value (PLV), which quantifies the consistency of the phase relationship between EEG signals from pairs of electrodes over time, indicating strength of synchronization. Further, proprietary features (US Patents No. 11,771,377 and No. 11,980,485) were employed. These features involved computing pairwise channel covariance matrices in well-established physiological frequency bands. In our nested models, feature selection was, in part, determined by mutual information.

### Model Development

Supervised prediction models were developed independently for several clinically meaningful outcomes: (i) Response to levetiracetam (LEV), using both unmedicated and medicated-state EEG recordings; (ii) Response prediction for lamotrigine (LTG) based on unmedicated EEG; (iii) Identification of patients who ultimately became treatment-refractory (non-responsive to multiple ASMs), using both unmedicated and medicated-state EEG recordings. Medicated-state EEG was defined as an EEG recording in which the individual was actively taking a prescribed ASM at the time of recording. Other classes of medication were not considered in this categorization. These models were trained separately for each clinical outcome using EEG-derived static and dynamic connectivity and entropy-based features.

### Validation Approach

Our primary method of assessing model performance was nested leave-one-out cross-validation (LOOCV). Under this approach, each individual is set aside once as the test case while the model is trained on all the remaining individuals. Within this training set, an additional inner loop is used to select which features to include and how many to keep, ensuring that these choices are made without ever seeing the held-out test case. This design prevents “data leakage” from the test set into the training process and provides an unbiased estimate of how the model would perform on new, unseen individuals. Additionally, LOOCV was chosen as opposed to k-fold techniques to maximize training size from the limited sample available for many of our models. We tested two machine learning architectures, architecture A and architecture B for each ASM model and present each of these results here.

### Statistical Analysis and Model Performance

Model performance was primarily measured by area under the receiver-operating-characteristic curve (AUC). For each model, we present two sets of sensitivity, specificity, and balanced accuracy results: one calculated based on a classification cut-off of 0.5 and another calculated post-hoc by optimizing for balanced accuracy. All machine learning analyses were executed in Python (Python version 3.10.5). All statistics were calculated using R (R version 4.5.0). Pairwise differences were assessed using two-tailed t-tests, with significance defined as *p*<0.05. Significance is presented before and after multiple testing correction with FDR. Effect sizes were determined via Cohen’s d, using pooled standard deviations. The code used for data preprocessing, feature extraction, and model development was version-controlled and archived to guarantee reproducibility and facilitate future analyses and refinements. The HEP dataset is not publicly downloadable; qualified investigators can request access from the HEP consortium (https://humanepilepsyproject.org) by submitting a data-use application and a project proposal that meets consortium approval.

## RESULTS

### Cohort Characteristics

We obtained EEG recordings with respective complete meta data annotations from 280 individuals (median age at recording 33 years; range 11-65 years; IQR 21.75-44.31) from 28 different hospital sites recorded from 5/12/2012 to 3/5/2018. An additional set of 59 EEGs was available but had incomplete annotations. Other available cohort characteristics are presented in **Table 1** prior to filtering for outliers. Individuals’ ages were rounded to the nearest year for deidentification purposes. A subset of 204 EEGs could also be matched to respective EEG clinical annotations containing information such as the presence of spikes, epileptiform activity, and seizures.

**Table 1.** Cohort characteristics.

| Variable | Overall<br>(n = 280) | LEV-unmed<br>(n = 61) | LEV-med<br>(n = 81) | LTG-unmed<br>(n = 40) | Refractory-<br>unmed<br>(n = 89) | Refractory-<br>med<br>(n = 122) |
| --- | --- | --- | --- | --- | --- | --- |
| Age at EEG, yr<br>median [IQR] | 33 [21.8-44.3] | 34.0 [23.0-47.0] | 34 [11.0-43.0] | 29.0 [18.8-44.3] | 33.0<br>[23.0-48.0] | 35.6 [23.3-44.0] |
| Number of<br>seizures during<br>enrollment, count<br>median [IQR] | 2 [0.0-21.5] | 0.0 [0.0-7.0] | 9 [1.8-49.3] | 7.5 [0.0-91.5] | 0.0 [0.0-87.3] | 3.0 [0.0-67.0] |
| Longest inter-seizure interval,<br>days median<br>[IQR] | 30 [10.0-85.0] | 46.0 [14.8-91.5] | 30 [7.8-90.0] | 14 [6.3-74.3] | 34.5 [13.3-87.3] | 65.3 [7.0-67.0] |
| Epileptiform<br>activity in EEG<br>present,<br>Yes/No** | 90/114 | 6/1 | 14/1 | 5/2 | 7/2 | 12/1 |
| Spikes in EEG<br>present,<br>Yes/No** | 29/147 | 2/5 | 4/11 | 2/5 | 3/6 | 3/10 |
| Slowing in EEG<br>present,<br>Yes/No** | 68/110 | 2/5 | 10/5 | 3/4 | 3/6 | 8/5 |
| Seizures during<br>EEG, Yes/No** | 31/165 | 6/0 | 14/0 | 5/0 | 7/0 | 12/0 |
| Dataset balance<br>after filters, ASM<br>responsive/ASM<br>resistant or non-responsive | 208/72 | 32/22 | 23/53 | 21/12 | 67/16 | 80/34 |

\*Some individuals met criteria for multiple cohorts
\*\*Some EEG annotations listed as “unknown” or not provided

### EEG Connectivity Signatures

As a preliminary investigation into the potential predictive ability of EEG features to differentiate responders and non-responders of particular ASM treatments, we examined a variety of static and dynamic connectivity metrics in different frequency ranges and across different response groups. This work revealed two important findings: 1) identification of novel dynamic features correlated with specific ASM outcomes and 2) distinct connectivity characteristics between EEGs recorded from individuals unmedicated and medicated on an ASM. We see some of these distinctions clearly on a group level in **Figure 1** with clearly increased dynamic connectivity across many electrode pairs in unmedicated non-responders to levetiracetam and lamotrigine (**Figure 1D** and **E**) compared to unmedicated responders (**Figure 1A** and **B**). These differences, however, are not immediately apparent in medicated responders and non-responders to levetiracetam (**Figure 1C** and **F**).

**Figure 1.**
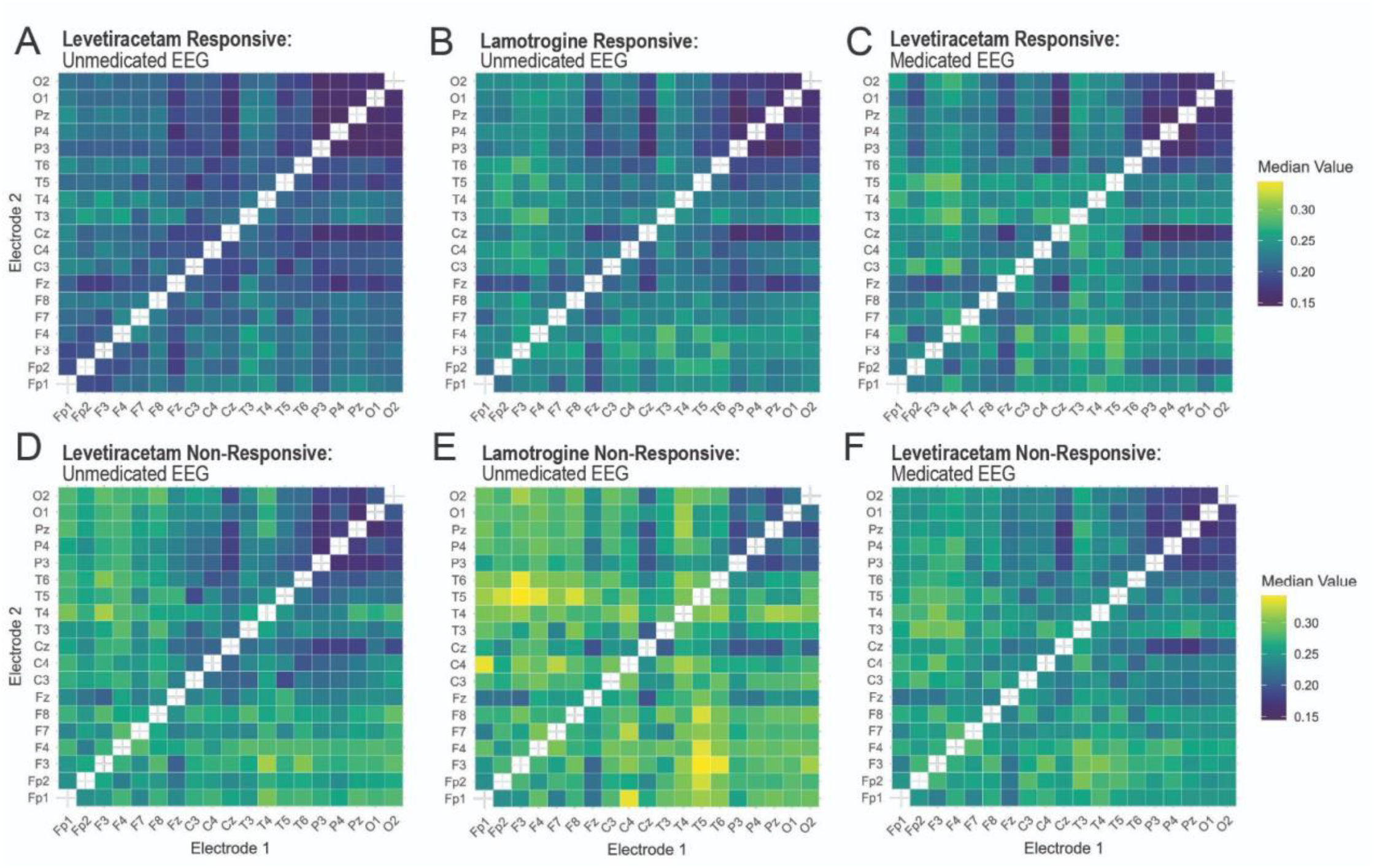
Novel dynamic connectivity feature demonstrating groupwise differences. A novel dynamic connectivity feature was calculated across EEGs at a high-frequency band. Each cell represents the median electrode pair value taken across individuals of responders and non-responders to levetiracetam (**A, C, D**, and **F**) and lamotrigine (**B** and **E**) who were in a medicated (**C** and **F**) and unmedicated (**A, B, D**, and **E**) state during their EEG. Colors were scaled to the minimum (blue) and max (yellow) value across groups.

These visual differences are supported statistically as well. Two-tailed t-tests within respective groups across electrodes reveal that unmedicated levetiracetam responders and non-responders have 36 significantly different electrode pairs with a mean effect size of 0.50 (95% CI: 0.46-0.53), 2 of which remain after multiple testing correction (mean effect size 0.75, 95% CI: 0.69-0.81). Unmedicated lamotrigine responders and non-responders have 20 significantly different electrode pairs with a mean effect size of 0.63 (95% CI: 0.59-0.66), 0 of which remain after multiple testing correction. Finally, medicated levetiracetam responders and non-responders have 83 significantly different electrode pairs with a smaller mean effect size of 0.34 (95% CI: 0.32-0.36), 58 of which remain after multiple testing correction (mean effect size 0.38, 95% CI: 0.36-0.39).

### Model Performance for Treatment Response

Our integration of diverse functional and entropy-based features into our multilayered model architectures resulted in strong performance across ASM and refractory prediction models, achieving AUCs on average of approximately 0.80 and above (**Table 2**; **Figure 2**). Models trained and tested on EEGs from individuals on no ASM at the time of recording reached higher performances than their respective medicated models. The top unmedicated levetiracetam model achieved a nested AUC of 0.88 and balanced accuracy of 0.85 at the optimized cut-off, while its medicated counterpart achieved an AUC 0.82 and balanced accuracy of 0.77 at the optimized cut-off. Similarly, the top unmedicated refractory model achieved a nested AUC of 0.92 and balanced accuracy of 0.87 at the optimized cut-off, while its medicated counterpart achieved an AUC of 0.82 and balanced accuracy of 0.75 at the optimized cut-off. Unmedicated lamotrigine performed worse, likely due to the small sample size, with its top prediction model architecture A achieving an AUC of 0.79 with a balanced accuracy of 0.80, sensitivity of 0.76, and specificity of 0.83 at an optimized cut-off of 0.65.

**Table 2.** ASM model nested LOOCV performance.

| Model | Architecture | AUC | Balanced Accuracy* | Sensitivity* | Specificity* |
| --- | --- | --- | --- | --- | --- |
| Unmedicated Levetiracetam | A | 0.88 | 0.85 (0.77) | 0.84 (0.90) | 0.86 (0.64) |
|  | B | 0.82 | 0.77 (0.69) | 0.81 (0.84) | 0.72 (0.55) |
| Medicated Levetiracetam | A | 0.81 | 0.80 (0.67) | 0.78 (0.39) | 0.81 (0.94) |
|  | B | 0.82 | 0.77 (0.71) | 0.70 (0.52) | 0.85 (0.91) |
| Unmedicated Lamotrigine | A | 0.79 | 0.80 (0.73) | 0.76 (0.95) | 0.83 (0.50) |
|  | B | 0.61 | 0.65 (0.59) | 0.71 (0.76) | 0.58 (0.42) |
| Unmedicated Refractory | A | 0.92 | 0.85 (0.63) | 0.70 (1.00) | 1.00 (0.25) |
|  | B | 0.92 | 0.87 (0.77) | 0.73 (0.97) | 1.00 (0.56) |
| Medicated Refractory | A | 0.81 | 0.76 (0.61) | 0.60 (0.86) | 0.91 (0.35) |
|  | B | 0.82 | 0.75 (0.72) | 0.68 (0.89) | 0.82 (0.56) |

\*Performance with optimized cut-off and performances with default 0.50 cut-off in parentheses

**Figure 2.**
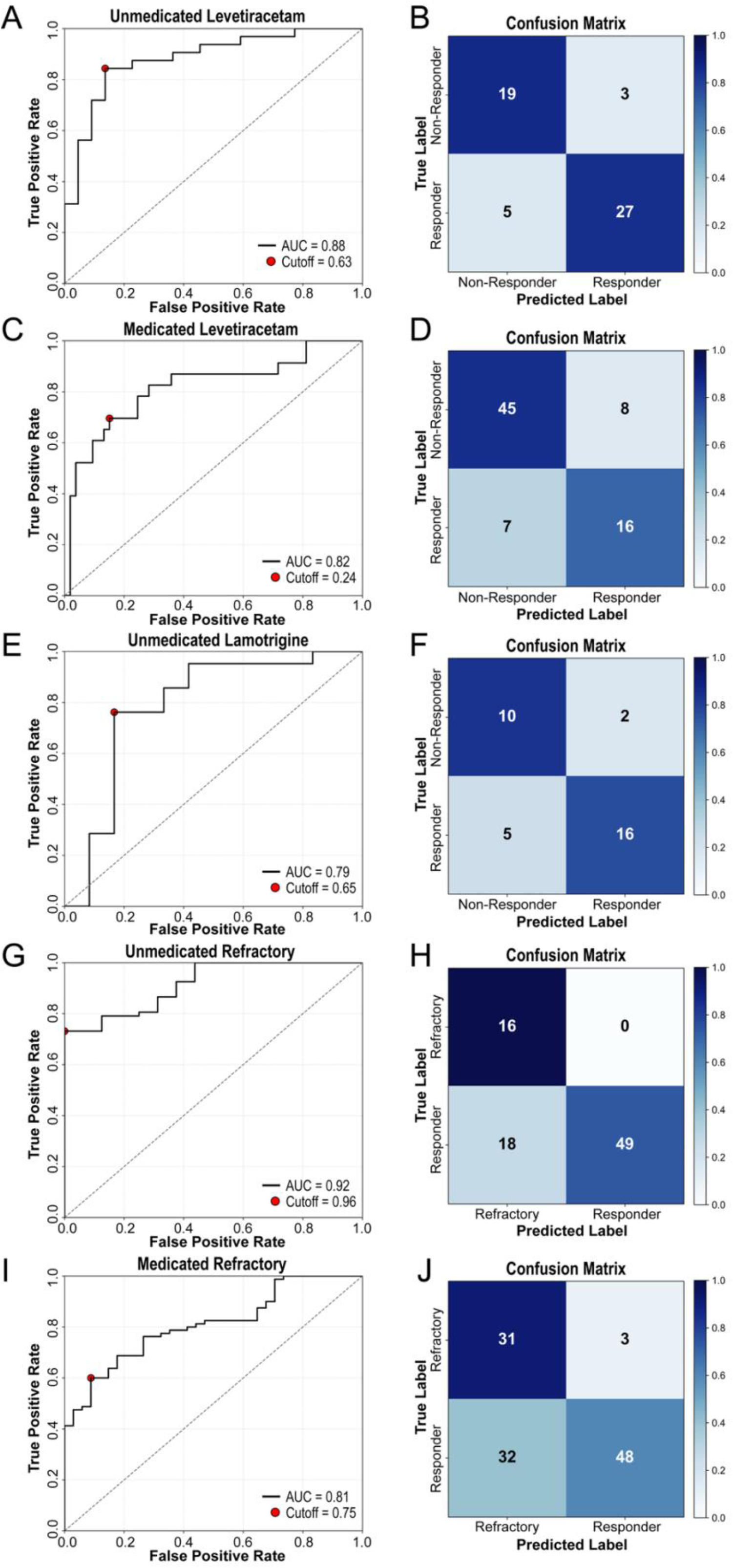
Nested cross-validation results. Receiver operator characteristic curves (ROC; left) and respective confusion matrices (right) resulting from the best-performing nested LOOCV model for each ASM model. Red dots indicate cut-offs determined post-hoc to result in the highest balanced accuracy. Confusion matrices results were determined by the respective model on the left and the calculated optimized cut-off. Shading indicates percentage of the respective class in either a true or false positive cell.

This poorer performance in EEGs recorded in a medicated state is likely due to a variety of factors; however, we hypothesized that a major contributor may be that the cohorts are on multiple different ASMs. To test this hypothesis, we underwent the same data preparation and model development for the medicated levetiracetam model, except only including individuals that were on levetiracetam at the time of the recording (n=53). This resulted in a drop in performance in both models, with its top model achieving an AUC of 0.75 with a balanced accuracy of 0.76, sensitivity of 0.74, and specificity of 0.79 at an optimized cut-off of 0.40. These findings suggest that the reduced performance cannot be explained solely by medication heterogeneity and that medication itself may alter EEG signals in ways that obscure predictive features.

### Exploratory Analysis on ASM Late Responders

The ILAE defines treatment refractory epilepsy as individuals who have tried and failed two or more appropriately dosed, tolerated ASMs [29]. A small percentage of individuals, however, go on to achieve seizure freedom on their third or later ASM. Due to this duality, we held these patients out when training and testing our refractory models. After filters, this held-out population consisted of 4 individuals unmedicated and 10 individuals medicated during their EEG. We hypothesized that these individuals’ electrophysiology has more in common with ILAE-defined treatment-responsive individuals than individuals non-responsive to all ASMs tried.

In the medicated population when applying a 0.5 cut-off, both Model A and Model B predicted 8/10 individuals to be responders. When applying a cut-off optimized on the training data, Model A and Model B predicted 8/10 and 7/10 individuals to be responders respectively. Both models predicted 4/4 unmedicated individuals to be responders when applying a 0.5 cut-off. When applying a cut-off optimized on the training data, Model A and Model B respectively predicted 1/4 and 3/4 individuals to be responders. Additional training and testing data is needed to make any concrete conclusions; however, the current preliminary results seem to support our hypothesis that treatment-refractory late-responder populations have an underlying electrophysiology more in common with patients responsive to their first or second ASM treatment than treatment-refractory patients who never achieve seizure freedom from an ASM.

## DISCUSSION

Accurately forecasting ASM response from a single baseline routine EEG has significant potential to revolutionize epilepsy management, moving beyond the trial-and-error approach commonly used today in the clinic to one guided by precision medicine. In this study, we introduce EEG-based machine learning algorithms demonstrating unprecedented predictive capabilities for individual patient responses to ASMs. Prior studies have not demonstrated that routine scalp EEG can accurately predict individual responses to lamotrigine. Our findings provide preliminary evidence that EEG-derived features can forecast lamotrigine and levetiracetam response and identify patients likely to develop drug-refractory epilepsy based on their baseline EEG. Additionally, our findings highlight a critical distinction between EEG biomarkers derived from unmedicated versus medicated states when predicting response to levetiracetam, suggesting that the presence of an ASM at the time of EEG recording influences predictive patterns. Our top models achieved high performances across all prediction tasks (AUC 0.79-0.92; **Figure 2**), demonstrating best-in-class accuracy that substantially surpasses prior solely EEG-based ASM response predictors [22, 32]. This represents a significant advancement toward precision medicine in epilepsy management, as our EEG biomarkers markedly outperformed existing approaches reliant on clinical features or conventional EEG analysis alone.

Beyond their technical performance, these EEG biomarkers carry important clinical and healthcare implications. Early selection of an effective ASM is crucial: non-optimal treatment in the initial stages of epilepsy has been associated with a poorer long-term prognosis [30, 33, 34]. Clinicians today face a proliferation of ASM options, yet the differences in efficacy among these drugs are often subtle and many ASMs overlap in their indications for focal and generalized seizures [35]. Each ASM also comes with a unique side-effect profile and variable effectiveness [36]; an ASM that controls seizures in one patient may fail or cause adverse effects in another. Previous attempts to predict medication response have largely relied on broad clinical characteristics of patients, without an in-depth, individualized analysis [18, 20, 37, 38]. In contrast, an objective biomarker-driven algorithm derived from EEG could guide the choice of therapy for a specific patient, potentially ensuring that the first medication has the highest probability of achieving seizure freedom. By minimizing the trial-and-error approach, such biomarkers could improve patient outcomes and reduce the healthcare costs associated with prolonged uncontrolled epilepsy and multiple medication trials.

Furthermore, being able to identify from the first EEG those patients unlikely to respond to any medication and those who have drug-resistant epilepsy (DRE) could prompt earlier consideration of alternative treatments such as epilepsy surgery or neurostimulation. From the time of initial epilepsy diagnosis, individuals that go on to receive resective or ablative surgery for DRE wait on average approximately 17 years in the United States before receiving their surgery [39]. This constitutes approximately two decades of failed ASM trials which not only do not control their seizures but frequently result in debilitating side-effects. Importantly, however, once a patient receives the additional diagnosis of DRE, the time to surgery is about 1-2 years [39]. As EEGs are typically recorded shortly before or after an epilepsy diagnosis, a validated and accurate EEG algorithm to predict DRE could shorten the time to surgery by more than a decade in some patients.

The impact of our model’s predictive power is evident when applied to levetiracetam and lamotrigine, two of the most commonly used ASMs. When levetiracetam or lamotrigine is chosen based on clinical judgment alone, the reported seizure freedom rates for levetiracetam vary as low as approximately 11-49% [40-44] and for lamotrigine ranges from 14-54% [44-46]. By contrast, our EEG model predicted lamotrigine and levetiracetam seizure freedom with 80-85% balanced accuracy in nested models. Such a tool could dramatically improve the odds of selecting the right patients for ASM monotherapy, avoiding ineffective trials often composed of side-effects, poor seizure control, higher healthcare costs, and poorer quality of life. This is especially beneficial for an agent such as lamotrigine which must be titrated slowly to avoid adverse effects, so an ineffective lamotrigine trial can leave a patient with uncontrolled seizures for months. Our model’s 80% accuracy in predicting lamotrigine response could help avoid such futile trials: clinicians could steer likely non-responders toward alternative therapies sooner and confidently use lamotrigine in those predicted to respond well. As our sample size grows, we anticipate the accuracy of our models to grow, particularly in our lamotrigine model which has the smallest sample size.

Our study significantly advances existing literature and compares favorably with recent EEG and genetics-based ASM prediction models. Prior EEG-based prediction studies often relied on smaller, single-center datasets with limited validation, raising concerns about generalizability and external validity; our rigorous validation approach addresses these issues, highlighting model stability and clinical applicability. Unlike Croce et al. (2021) and Zhang et al. (2018), who specifically targeted levetiracetam monotherapy and achieved moderate predictive performance (AUC range: 0.61–0.84) using EEG spectral entropy features [22, 23], our study incorporates advanced functional connectivity EEG features, achieving superior performance (AUC up to 0.92 in nested models). Wang et al. (2022) similarly employed EEG complexity metrics, but our broader medication focus encompassing levetiracetam, lamotrigine, and refractory epilepsy prediction and robust validation methods provide stronger clinical relevance [24]. Compared to genetic-based predictions by de Jong et al. (2021) which achieved an AUC of 0.75 [25], our EEG-based approach offers considerable practical advantages, as routine EEG recordings are already standard in epilepsy care. In contrast, comprehensive genomic analyses, despite their theoretical potential, remain costly, less efficient, and clinically impractical for routine use. Thus, our methodology uniquely combines state-of-the-art predictive accuracy with practical clinical feasibility, enabling immediate translational potential. Nevertheless, future studies incorporating multimodal data, such as combining EEG with syndromic and genetic information when feasible, will be important to further enhance prediction accuracy and advance precision epilepsy management.

Methodologically, our approach leverages a robust, data-driven machine-learning pipeline, previously validated in antidepressant and placebo-response prediction for major depressive disorder [28]. In earlier work, we successfully identified distinct EEG subtypes predicting response to SSRIs versus placebo in MDD, underscoring the reproducibility and predictive strength of our methodology across neuropsychiatric conditions [28]. Thus, the high predictive performance achieved here in epilepsy further validates our pipeline’s utility for precision neuropsychiatry.

Despite these methodological strengths, certain limitations warrant consideration. Firstly, our analysis utilized retrospective data from a single multicenter cohort, limiting immediate generalizability. The modest sample size, especially within certain subgroups such as lamotrigine responders, underscores the necessity for larger, prospective validation studies. Moreover, epilepsy encompasses an extremely diverse set of subtypes with a variety of seizure types, comorbidities, and, in some cases, distinct electrophysiology [47, 48]. The HEP cohort used here encompasses one, albeit large, segment of the epilepsy population: generally healthy patients with new-onset focal epilepsy with normal IQ and without recent large brain lesions. Furthermore, we evaluated only two ASMs, whereas clinicians frequently face numerous treatment options. Prospective extensions to additional ASMs and drug combinations are necessary to maximize clinical applicability. Lastly, potential confounders such as variability in EEG timing relative to medication initiation, ASM dosage, erroneous seizure frequency annotations, lower sampling rate, and patient heterogeneity may have influenced predictive performance, warranting careful consideration in future studies. We have already begun to gather and annotate thousands of additional retrospective EEGs across multiple independent cohorts from different hospital systems throughout the United States. With this extensive and growing dataset, we aim to validate and extend our models’ predictive capabilities to current and additional ASMs.

In conclusion, our study demonstrates that routine interictal EEG analyzed using advanced machine learning can reliably forecast individual patient responses to specific ASMs and predict medically refractory epilepsy. By enabling earlier selection of effective treatments, these EEG biomarkers could fundamentally transform epilepsy management, aligning epilepsy care with the broader goals of precision medicine. Collectively, these findings represent a substantial step toward personalized epilepsy therapy and support further integration of EEG biomarkers into clinical practice.

## Data Availability

All data analyzed in the present study originates from the Human Epilepsy Project (HEP#1) dataset which is not publicly downloadable. Qualified investigators can request access from the HEP consortium (https://humanepilepsyproject.org) by submitting a data-use application and a project proposal that meets consortium approval.

## REFERENCES

1. Kobau, R., C. Luncheon, and K. Greenlund, Active epilepsy prevalence among U.S. adults is 1.1% and differs by educational level—National Health Interview Survey, United States, 2021. Epilepsy & Behavior, 2023. 142: p. 109180.

2. Chen, Z., et al., Editorial: Epidemiology of epilepsy and seizures. Front Epidemiol, 2023. 3: p. 1273163.

3. Sperling, M.R., et al., Seizure control and mortality in epilepsy. Ann Neurol, 1999. 46(1): p. 45–50.

4. Foster, E., et al., Comparisons of direct and indirect utilities in adult epilepsy populations: A systematic review. Epilepsia, 2019. 60(12): p. 2466–2476.

5. Begley, C., et al., The global cost of epilepsy: A systematic review and extrapolation. Epilepsia, 2022. 63(4): p. 892–903.

6. Allers, K., et al., The economic impact of epilepsy: a systematic review. BMC Neurology, 2015. 15(1): p. 245.

7. Pallin, D.J., et al., Seizure visits in US emergency departments: epidemiology and potential disparities in care. Int J Emerg Med, 2008. 1(2): p. 97–105.

8. Moura, L., I. Karakis, and D. Howard, Emergency department utilization among adults with epilepsy: A multi-state cross-sectional analysis, 2010–2019. Epilepsy Research, 2024. 205: p. 107427.

9. Yu, Y., et al., Nationwide Patterns of Healthcare Utilization and Health related Quality of Life among US Adults with Epilepsy (P6-1.002). Neurology, 2023. 100(17_supplement_2): p. 3385.

10. Cramer, J.A., et al., Healthcare utilization and costs in adults with stable and uncontrolled epilepsy. Epilepsy Behav, 2014. 31: p. 356–62.

11. Chen, Z., et al., Treatment Outcomes in Patients With Newly Diagnosed Epilepsy Treated With Established and New Antiepileptic Drugs: A 30-Year Longitudinal Cohort Study. JAMA Neurol, 2018. 75(3): p. 279–286.

12. Pisani, L.R., et al., Specific Patient Features Affect Antiepileptic Drug Therapy Decisions: Focus on Gender, Age, and Psychiatric Comorbidities. Curr Pharm Des, 2017. 23(37): p. 5639–5648.

13. Beniczky, S., et al., Optimal choice of antiseizure medication: Agreement among experts and validation of a web-based decision support application. Epilepsia, 2021. 62(1): p. 220–227.

14. Galer, P.D., et al., Quantitative EEG Biomarkers in the Genetic Epilepsies and Associations With Neurologic Outcomes. Neurology, 2025. 105(8): p. e214148.

15. Myers, P., et al., Diagnosing Epilepsy with Normal Interictal EEG Using Dynamic Network Models. Ann Neurol, 2025. 97(5): p. 907–918.

16. Yao, L., et al., Prediction of antiepileptic drug treatment outcomes of patients with newly diagnosed epilepsy by machine learning. Epilepsy & Behavior, 2019. 96: p. 92–97.

17. Yang, S., B. Wang, and X. Han, Models for predicting treatment efficacy of antiepileptic drugs and prognosis of treatment withdrawal in epilepsy patients. Acta Epileptologica, 2021. 3(1): p. 1.

18. Devinsky, O., et al., Changing the approach to treatment choice in epilepsy using big data. Epilepsy Behav, 2016. 56: p. 32–7.

19. An, S., et al., Predicting drug-resistant epilepsy - A machine learning approach based on administrative claims data. Epilepsy Behav, 2018. 89: p. 118–125.

20. Hakeem, H., et al., Development and Validation of a Deep Learning Model for Predicting Treatment Response in Patients With Newly Diagnosed Epilepsy. JAMA Neurology, 2022. 79(10): p. 986–996.

21. Smolyansky, E.D., et al., Machine learning models for decision support in epilepsy management: A critical review. Epilepsy & Behavior, 2021. 123: p. 108273.

22. Zhang, J.H., et al., Personalized prediction model for seizure-free epilepsy with levetiracetam therapy: a retrospective data analysis using support vector machine. Br J Clin Pharmacol, 2018. 84(11): p. 2615–2624.

23. Croce, P., et al., Machine learning for predicting levetiracetam treatment response in temporal lobe epilepsy. Clinical Neurophysiology, 2021. 132(12): p. 3035–3042.

24. Wang, B., et al., EEG-Driven Prediction Model of Oxcarbazepine Treatment Outcomes in Patients With Newly-Diagnosed Focal Epilepsy. Frontiers in Medicine, 2022. 8.

25. de Jong, J., et al., Towards realizing the vision of precision medicine: AI based prediction of clinical drug response. Brain, 2021. 144(6): p. 1738–1750.

26. The Human Epilepsy Project, C. The Epilepsy Study, Editor. 2014.

27. Fox, J., et al., Patterns of antiseizure medication utilization in the Human Epilepsy Project. Epilepsia, 2023. 64(12): p. 3196–3204.

28. Li, Q., et al., Machine Learning-Enabled EEG Biomarkers Predict Divergent Antidepressant and Placebo Response in a Clinical Trial of Major Depression. medRxiv, 2025: p. 2025.05.29.25328167.

29. Kwan, P., et al., Definition of drug resistant epilepsy: consensus proposal by the ad hoc Task Force of the ILAE Commission on Therapeutic Strategies. Epilepsia, 2010. 51(6): p. 1069–77.

30. Kwan, P. and M.J. Brodie, Early identification of refractory epilepsy. N Engl J Med, 2000. 342(5): p. 314–9.

31. Saby, J.N., et al., Electrophysiological biomarkers of brain function in CDKL5 deficiency disorder. Brain Commun, 2022. 4(4): p. fcac197.

32. Croce, P., et al., Machine learning for predicting levetiracetam treatment response in temporal lobe epilepsy. Clin Neurophysiol, 2021. 132(12): p. 3035–3042.

33. Bjørke, A.B., et al., Evaluation of long-term antiepileptic drug use in patients with temporal lobe epilepsy: Assessment of risk factors for drug resistance and polypharmacy. Seizure, 2018. 61: p. 63–70.

34. Pellinen, J., J. French, and K.G. Knupp, Diagnostic Delay in Epilepsy: the Scope of the Problem. Curr Neurol Neurosci Rep, 2021. 21(12): p. 71.

35. Löscher, W. and P. Klein, The Pharmacology and Clinical Efficacy of Antiseizure Medications: From Bromide Salts to Cenobamate and Beyond. CNS Drugs, 2021. 35(9): p. 935–963.

36. Mao, J., et al., Real-world anti-seizure treatment and adverse events among individuals living with drug-resistant focal epilepsy in the United States. Epilepsia Open, 2024. 9(4): p. 1311–1320.

37. Abdaltawab, A., et al., How accurate are machine learning models in predicting anti-seizure medication responses: A systematic review. Epilepsy & Behavior, 2025. 163: p. 110212.

38. Delen, D., et al., Using predictive analytics to identify drug-resistant epilepsy patients. Health Informatics J, 2020. 26(1): p. 449–460.

39. Campbell, J.M., et al., Delays in the diagnosis and surgical treatment of drug-resistant epilepsy: A cohort study. Epilepsia, 2024. 65(5): p. 1314–1321.

40. Stephen, L.J., et al., Levetiracetam monotherapy—Outcomes from an epilepsy clinic. Seizure, 2011. 20(7): p. 554–557.

41. Berkovic, S.F., et al., Placebo-controlled study of levetiracetam in idiopathic generalized epilepsy. Neurology, 2007. 69(18): p. 1751–1760.

42. Depondt, C., et al., The long term retention of levetiracetam in a large cohort of patients with epilepsy. J Neurol Neurosurg Psychiatry, 2006. 77(1): p. 101–3.

43. Devinsky, O. and C. Elger, Efficacy of levetiracetam in partial seizures. Epileptic Disord, 2003. 5 Suppl 1: p. S27–31.

44. Arif, H., et al., Comparative effectiveness of 10 antiepileptic drugs in older adults with epilepsy. Arch Neurol, 2010. 67(4): p. 408–15.

45. Rosenow, F., et al., The LaLiMo Trial: lamotrigine compared with levetiracetam in the initial 26 weeks of monotherapy for focal and generalised epilepsy--an open-label, prospective, randomised controlled multicenter study. J Neurol Neurosurg Psychiatry, 2012. 83(11): p. 1093–8.

46. Knoester, P.D., et al., Effectiveness of lamotrigine in clinical practice: Results of a retrospective population-based study. Epilepsy Research, 2005. 65(1): p. 93–100.

47. Seneviratne, U., M.J. Cook, and W.J. D’Souza, Electroencephalography in the Diagnosis of Genetic Generalized Epilepsy Syndromes. Front Neurol, 2017. 8: p. 499.

48. Wirrell, E.C., et al., Methodology for classification and definition of epilepsy syndromes with list of syndromes: Report of the ILAE Task Force on Nosology and Definitions. Epilepsia, 2022. 63(6): p. 1333–1348.

